# Machine learning for the prediction of in-hospital mortality in patients with spontaneous intracerebral hemorrhage

**DOI:** 10.1101/2023.08.15.23294147

**Authors:** Baojie Mao, Rui Zhang, Yuhang Pan, Ruzi Zheng, Yanfei Shen, Wei Lu, Yuning Lu, Shanhu Xu, Jiong Wu, Ming Wang, Shu Wan

## Abstract

**Backgrounds:** Early and accurate identification of patients with spontaneous intracerebral hemorrhage(sICH) who are at high risk of in-hospital death can help intensive care unit (ICU) physicians make optimal clinical decisions. The aim of this study was to develop a machine learning(ML)-based tool to predict the risk of in-hospital death in patients with sICH in ICU.

**Methods:** We conducted a retrospective administrative database study using the MIMIC-IV and Zhejiang Hospital database. The outcome of the study was in-hospital mortality. To develop and validate the final model, we employed the LASSO regression to screen and select relevant variables. Five algorithms, namely Logistic Regression (LR), K-Nearest Neighbors (KNN), Adaptive Boosting (AdaBoost), Random Forest (RF), and eXtreme Gradient Boosting (XGBoost), were utilized. The selection of the best model was based on the area under the curve (AUC) in the validation cohort. Furthermore, we employ the SHapley Additive exPlanations (SHAP) methodology to elucidate the contributions of individual features to the model and analyze their impact on the model’s outputs. To facilitate accessibility, we also created a visual online calculation page for the model.

**Results:** In the final cohort comprising 1596 patients from MIMIC-IV and Zhejiang Hospital, 367 individuals (23%) experienced in-hospital mortality during the inpatient follow-up period. After extracting 46 variables from the database, LASSO regression identified 14 predictor variables for further analysis. Among the five evaluated models, the XGBoost model demonstrated superior discriminative power in both the internal validation set (AUC = 0.907) and the external validation set (AUC = 0.787). Furthermore, through the SHAP technique, we identified the top 5 predictors in the feature importance rankings: Glasgow Coma Scale (GCS), Sequential Organ Failure Assessment (SOFA), anticoagulant medication, mannitol medication and oxygen saturation.

**Conclusions:** Among the five models, the XGBoost model exhibited superior performance in predicting mortality for patients with sICH in the ICU, indicating its potential significance in the development of early warning systems.

---

Spontaneous intracerebral hemorrhage (sICH) is a critical neurological event characterized by bleeding within the intracerebral parenchyma, leading to a sudden and potentially life-threatening medical emergency^1^. Despite notable advancements in medical care, the overall prognosis of sICH remains poor, primarily attributed to inflammatory responses, oxidative stress, and other mechanisms, resulting in a significant proportion of patients succumbing during hospitalization ^2,3^. Accurate prediction of in-hospital mortality holds paramount importance in guiding clinical decision-making and optimizing resource allocation. Therefore, there is an urgent need for an advanced model that can help predict the prognosis of patients with sICH.

In recent years, there has been a growing interest in the application of machine learning (ML) techniques to investigate various clinical diseases ^4–8^. Compared to traditional statistics, ML exhibits characteristics such as handling complex nonlinear data, offering high flexibility, and enabling continuous learning and improvement ^9,10^. Prior attempts have been made in the field of sICH ^11,12^. However, there is still a pressing need for models that can accurately predict severe sICH cases in ICU.

In this study, our objective is to develop and validate a machine learning-based predictive model for in-hospital mortality among sICH patients. By utilizing a comprehensive set of clinical and demographic features, we aim to provide clinicians with a robust tool to accurately assess the risk of mortality in real-time, thus facilitating timely interventions and improving patient care.

## Database

The Medical Information Mart for Intensive Care-IV (MIMIC-IV) is an open and freely accessible critical care database that contains comprehensive clinical data of patients admitted to a tertiary academic medical center in Boston, MA, USA, from 2008 to 2019. The database includes essential patient information, vital signs, laboratory indicators, treatment details, and survival data. The usage of data from MIMIC-IV has been granted approval by the Institutional Review Boards of Beth Israel Deaconess Medical Center (Boston, MA) and Massachusetts Institute of Technology (MIT; Cambridge, MA). As all personal data in this database is encrypted, informed consent was waived. One of the authors (Mao, Baojie) obtained access to the database and was responsible for data extraction (certification number 46148427). In addition, we recruited patients with cerebral hemorrhage who were admitted to ICU from December 2018 to February 2023 in Zhejiang Hospital. The study protocol was approved by the Ethics Committee of Zhejiang Hospital (No. 2023 Pro-examination (58K)). All patient data were anonymized. No patient-identifiable data were recorded throughout the study. Given that this study was purely observational, written consent from patients was not required.

### Data extraction and outcomes

Clinical and laboratory variables were meticulously collected within 24 hours of admission to the Intensive Care Unit (ICU). In the case of variables with multiple measurements, mean values were calculated and utilized for analysis. A total of forty-six variables were included in the data collection process. These encompassed patient characteristics (age, gender), vital signs (respiratory rate, blood pressure, heart rate, oxygen saturation, and temperature), laboratory data (routine blood analysis, renal function, coagulation, and blood gases), as well as comorbidities identified based on recorded International Classification of Diseases ICD-9 and ICD-10 codes. The comorbidities considered were hypertension, diabetes mellitus, chronic obstructive pulmonary disease (COPD), congestive heart failure, renal disease, liver disease, and malignancy. Furthermore, information regarding the usage of anticoagulant and vasoactive drugs, surgical status, Glasgow Coma Scale (GCS), Sequential Organ Failure Assessment (SOFA) scores, mechanical ventilation, and renal replacement therapy (RRT) was gathered. Due to the limited number of patients with missing data, we opted to exclude them from the analysis rather than attempting to estimate the missing values.The primary endpoint was all-cause in-hospital mortality.

### Cohort selection

1. Patients must be admitted to the ICU for the first time.
2. Patients must have a confirmed diagnosis of sICH.
3. Patients’ age should fall within the range of 18 to 90 years.
4. Patients must have an ICU length of stay exceeding 1 day.
5. Patients must have complete clinical data.

The flowchart for patient recruitment is shown in Figure 1.

**Figure 1.**
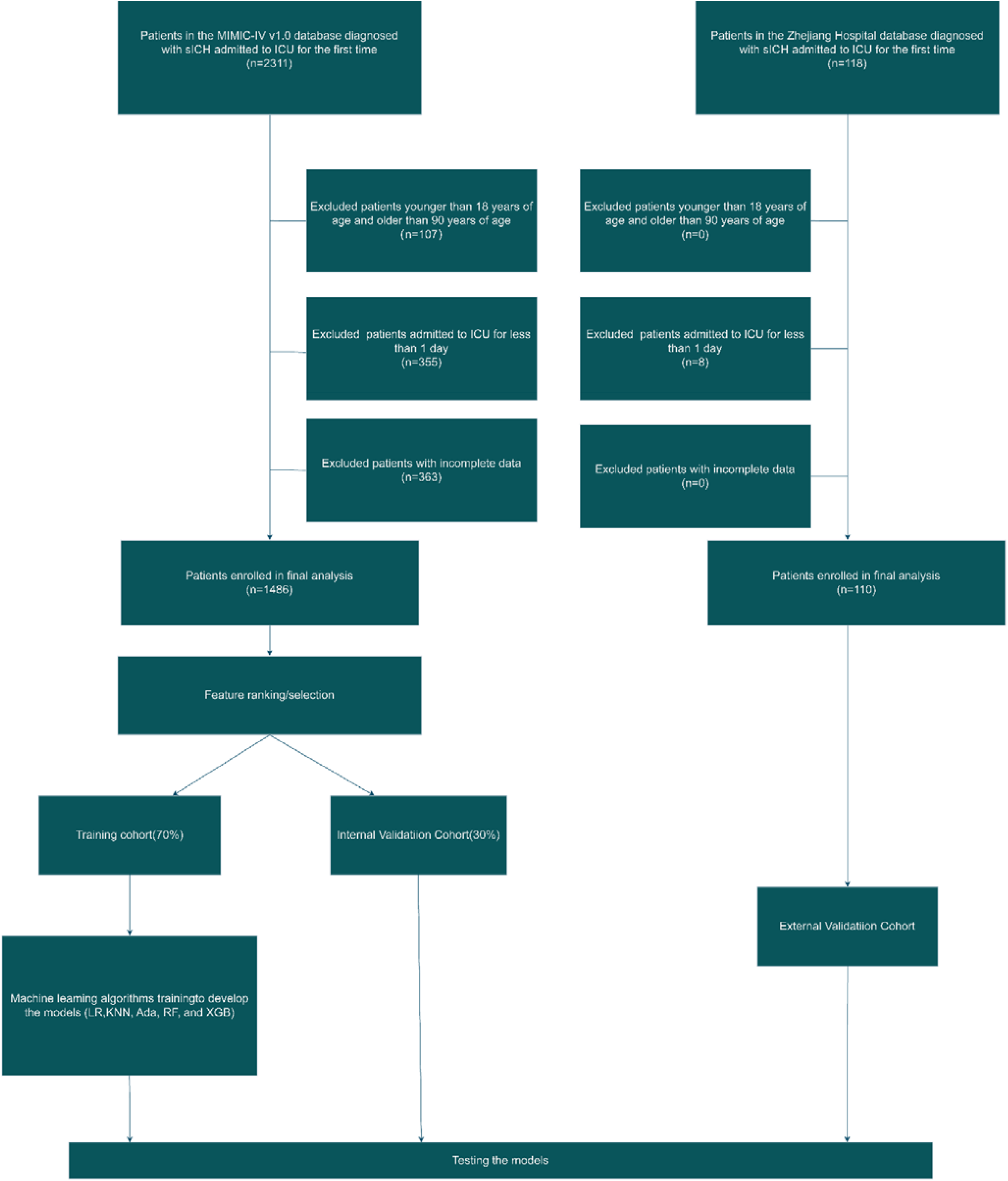
Model development process and flowchart of the study

**Figure 2.**
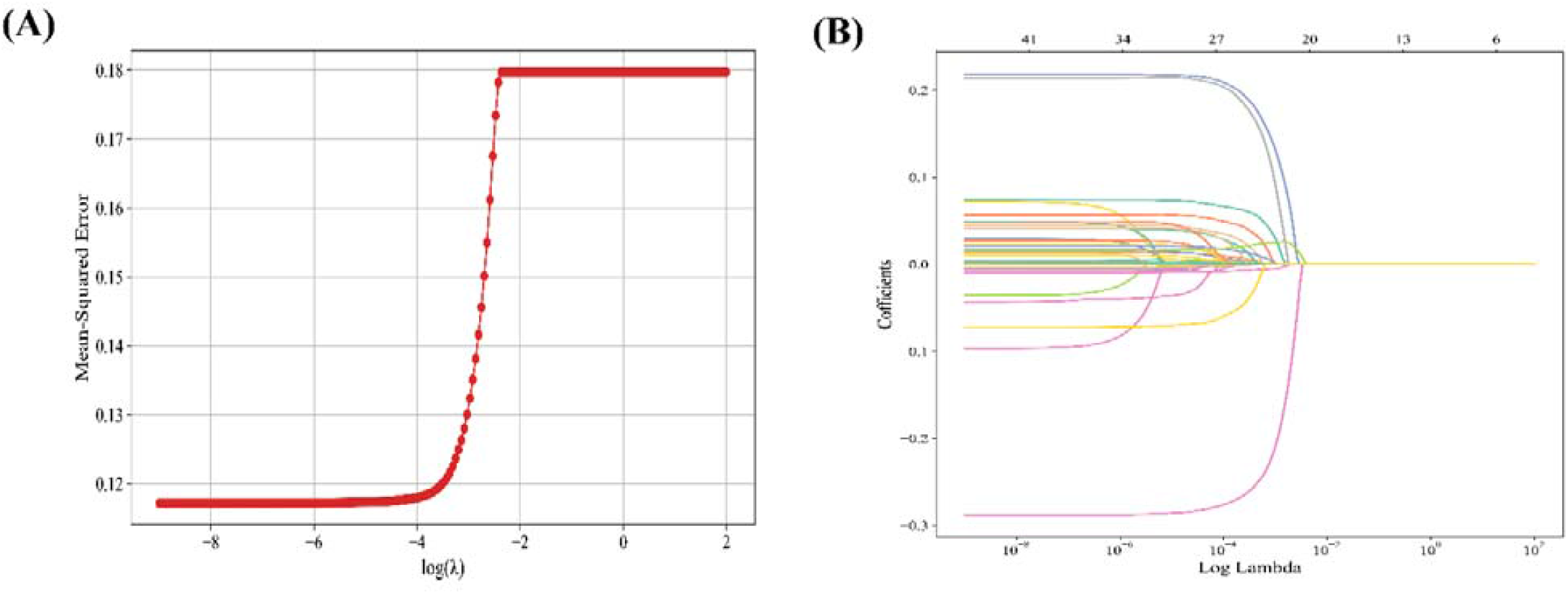
Demographic and clinical feature selection using the LASSO regression

### Feature Selection

We applied Lasso regression, a regularization technique, on the preprocessed dataset. Lasso performs feature selection by shrinking the coefficients of less important features to zero, effectively eliminating them from the model. The optimal regularization parameter (λ) for Lasso was determined using the coordinate descent algorithm. Following Lasso regression, the variables were ranked based on their corresponding non-zero coefficients. The final predictive model included the top 14 variables with the highest absolute coefficient values.

### Statistical analysis

The normality of the distribution was evaluated using the Kolmogorov-Smirnov test. Continuous variables were presented as mean with standard deviation if they followed a normal distribution, or as median with 25th to 75th percentile if they deviated from normality. The Student’s t-test or Mann-Whitney test was applied accordingly to analyze the continuous variables. Categorical variables were presented as counts and percentages, and the chi-square test was utilized to compare the distributions.

In this study, we employed five different ML algorithms to develop models: Logistic Regression (LR), K-Nearest Neighbors (KNN), Adaptive Boosting (AdaBoost), Random Forest (RF) and eXtreme Gradient Boosting algorithms (XGBoost). The MIMIC IV dataset was initially partitioned into a training set (70%) and an internal validation set (30%). Furthermore, we utilized the Zhejiang Hospital dataset as an external validation set. The validation process employed a bootstrap resampling technique with 1000 iterations to evaluate the model’s performance. The Area Under the Curve (AUC) and 95% confidence intervals (CI) were calculated. Furthermore, several evaluation metrics, including accuracy, sensitivity, specificity, Youden index, and F1 score, were computed. For hyperparameter selection, 5-fold cross-validation and grid search methods were utilized.

To assess the performance and clinical applicability of the predictive model, we generated calibration curves and clinical decision curves. Calibration curves were used to evaluate the predictive accuracy and calibration of the model by comparing the predicted probabilities with actual observations. On the other hand, clinical decision curves were employed to determine the model’s sensitivity and specificity at various decision thresholds, thus optimizing its predictive performance for clinical decision-making. After selecting the optimal model, we utilized the Shapley Additive exPlanations (SHAP) package in Python to demonstrate the importance of each feature. Subsequently, we employed the selected model to visualize prospective validations. Statistical significance was set at P < 0.05, and all tests were two-tailed. Statistical analyses were performed using R software (version 4.3.1) or Python software (version 3.11).

## Result

### Baseline Characteristics

The present study involved a total of 1596 patients, including 1486 patients from the internal cohort extracted from the MIMIC-IV database and 110 patients from the external cohort extracted from the Zhejiang Hospital database. In the internal cohort, there were 349 in-hospital deaths (23.48%), whereas the external cohort had 18 in-hospital deaths (16.36%). Table 1 provides an overview of the baseline characteristics for both the internal and external cohorts.

**Table 1.**
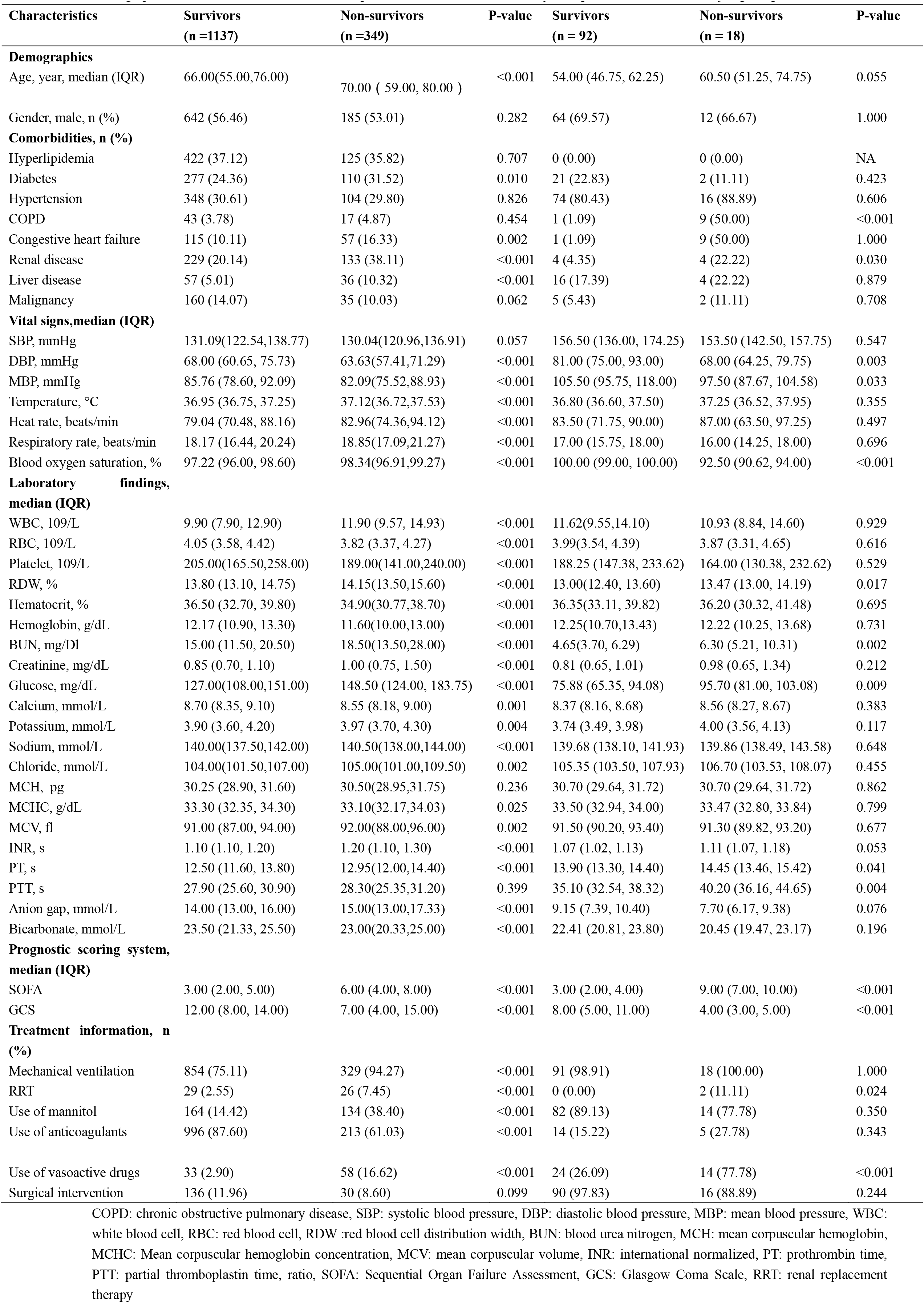
Demographic and Clinical Characteristics of Hospitalization Survival and Mortality Groups in MIMIC IV and Zhejiang Hospital Database.

### Key variables

This study utilized Lasso regression for feature selection by determining an appropriate lambda value. From the initial pool of 46 candidate variables, we identified the top 14 based on their importance and integrated them into the final model. The selected variables encompassed: use of anticoagulants, use of mannitol, use of vasoactive drugs, mechanical ventilation, temperature, surgical intervention, serum potassium, heart failure, blood oxygen saturation, SOFA, GCS, serum sodium, RDW and serum chloride.

### Model performance

The discriminative ability of all models to predict mortality is shown in Figure 3 and Table 2. In the training set, XGBoost, KNN, LR, RF, and AdaBoost models were built, and the AUCs of the internal validation set were 0. 907, 0.808, 0.851, 0.897, and 0.900, respectively. In contrast, the prediction performance of the XGBOOST model was the highest among these five models (AUC 0.907; 95% CI: 0.875 -0.939; accuracy: 0.874; sensitivity: 0.582). In the external validation set, the XGBoost model demonstrated excellent predictive power with an AUC of 0.788, second only to the LR model, which achieved an AUC of 0.790.

**Figure 3.**
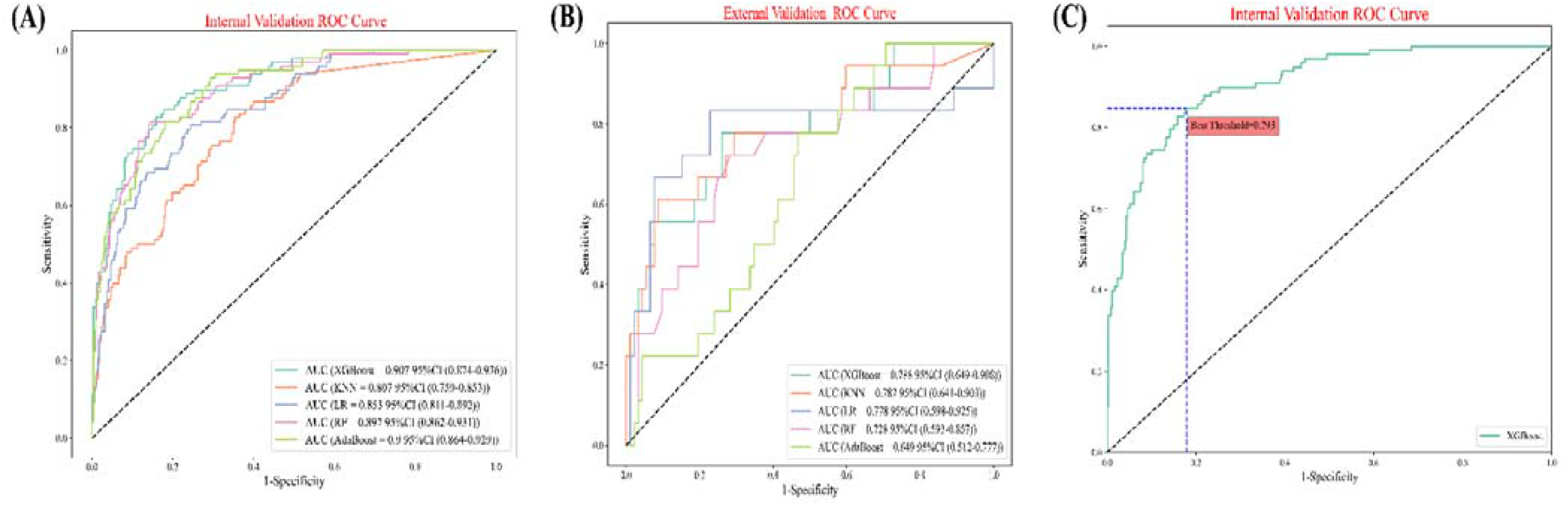
Area under the receiver operating characteristic curve for machine learning models in the internal validation queue and external validation queue. ROC: receiver operate characteristics, CI: confidence intervals

**Table 2.** Predictive performance of machine learning models in internal and external validation sets.

| Model | AUC | Accuracy | Sensitivity | Specificity | Youden Index | F1 score |
| --- | --- | --- | --- | --- | --- | --- |
| Internal validation cohort |  |  |  |  |  |  |
| XGBoost | 0.907 | 0.874 | 0.582 | 0.957 | 0.769 | 0.671 |
| KNN | 0.807 | 0.823 | 0.337 | 0.960 | 0.648 | 0.445 |
| Logistic Regression | 0.851 | 0.843 | 0.551 | 0.925 | 0.738 | 0.606 |
| Random Forest | 0.897 | 0.858 | 0.602 | 0.931 | 0.767 | 0.652 |
| AdaBoost | 0.900 | 0.865 | 0.561 | 0.951 | 0.756 | 0.647 |
| External validation cohort |  |  |  |  |  |  |
| XGBoost | 0.788 | 0.554 | 0.833 | 0.500 | 0.667 | 0.380 |
| KNN | 0.787 | 0.855 | 0.500 | 0.924 | 0.712 | 0.529 |
| Logistic Regression | 0.790 | 0.436 | 0.833 | 0.359 | 0.596 | 0.326 |
| Random Forest | 0.728 | 0.436 | 0.833 | 0.359 | 0.596 | 0.326 |
| AdaBoost | 0.650 | 0.500 | 0.778 | 0.446 | 0.612 | 0.337 |

The optimal cut-off for prediction probabilities of the XGBoost model was determined to be 29.53% based on the Youden Index, which is calculated as sensitivity + specificity -1(Figure 3C).

Figure 4A shows the calibration plots for all five models. The calibration curve analysis showed that XGBoost was accurately calibrated in predicting the risk of in-hospital death, with no significant over or underestimation(Figure 4B). In addition, the results of the Decision Curve Analysis (DCA) clearly support the use of the XGBoost model as a valid predictive tool for patients with severe sICH, as shown in Figure 4C and 4D.

**Figure 4.**
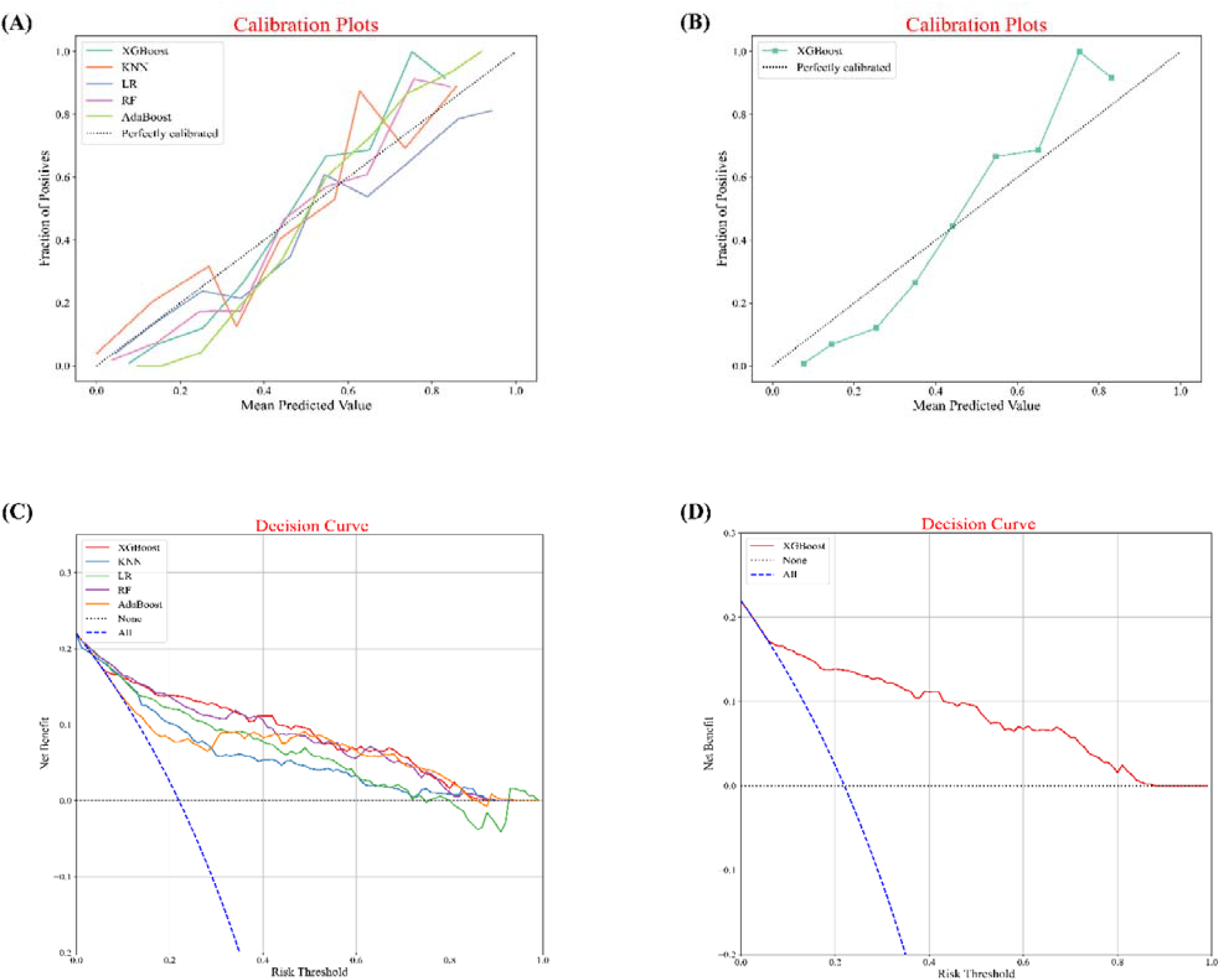
Calibration plots of five models (A, B). XGBOOST has no significant bias in model predictions. Decision curve analysis for five machine learning models(C, D). The XGBoost model represents an ideal predictive tool for forecasting in-hospital mortality.

Feature importance analysis was performed to identify the important features that influence the model output. The importance of features derived from XGBoost model is shown in Figure 5.GCS score was the most influential feature followed by SOFA score, use of anticoagulants, use of mannitol, oxygen saturation, body temperature, serum sodium, serum potassium, RDW, mechanical ventilation, heart failure, serum chloride, use of vasoactive drugs and surgical intervention.

**Figure 5.**
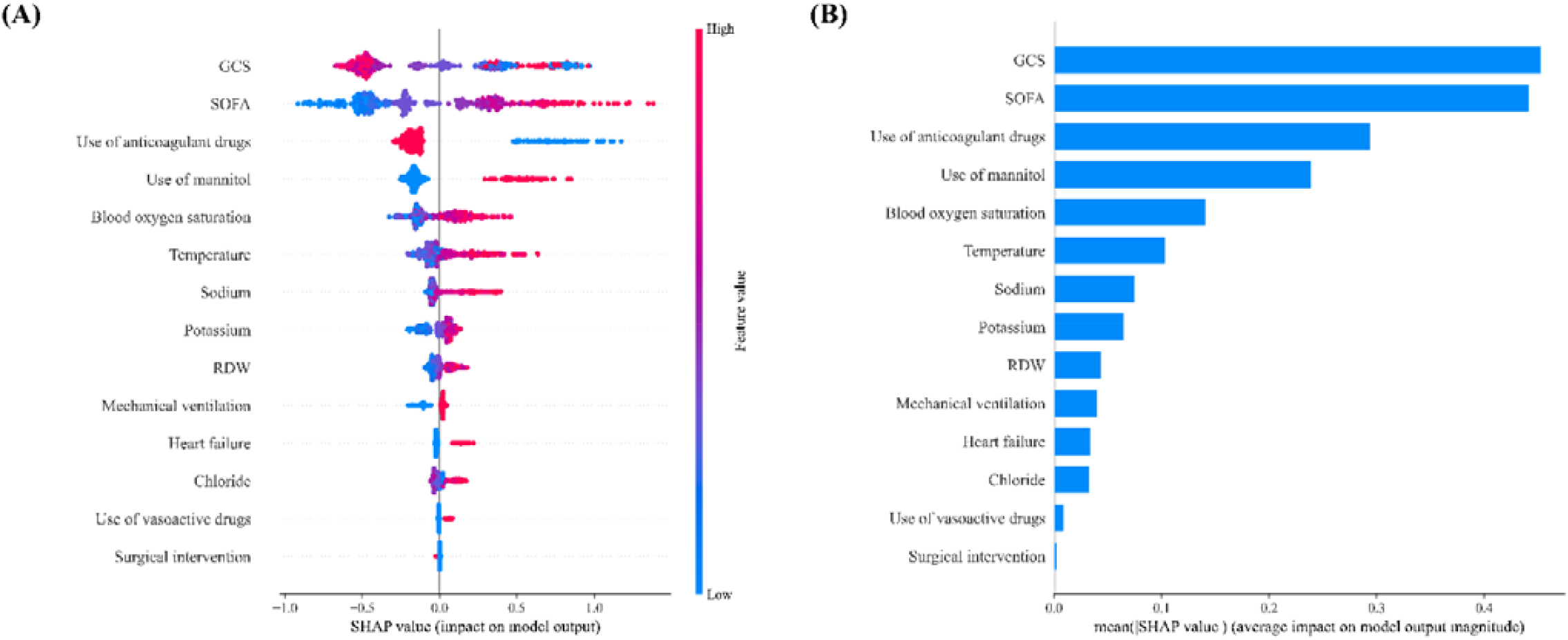
SHAP analysis of the XGBoost model. A visual representation illustrates the importance of each feature in the XGBoost model, depicting the relationship between them. The color scale indicates the variable values, with red denoting higher values and blue indicating lower values.

### Application of the model

Additionally, a web-based computational tool has been developed to enable clinicians in real-time prediction of the prognosis for patients with severe sICH. (accessible at https://ich-wpp0azhoyz.streamlit.app/) Clinical information of a typical patient was entered into the model, for example, use of anticoagulants: no, use of mannitol: no, use of vasoactive drugs: no, mechanical ventilation: yes, heart failure: no, surgical intervention: no, serum chlorine: 108 mmol/L, GCS score: 5, SOFA score: 8, temperature: 37.6°C, RDW: 13%, serum sodium: 146 mmol/ L; serum potassium: 3.7 mmol/L; oxygen saturation: 99%. The model predicted that this patient had a 70.846% risk of in-hospital mortality, indicating that this patient was at high risk and that medical staff should be prepared to treat and care for him in advance (Figure 6A). Information from another patient was included in the model: use of anticoagulants: no, use of mannitol: no, use of vasoactive medications: yes, mechanical ventilation: yes, heart failure: no, surgical intervention: yes, serum chloride: 102 mmol/L, GCS score: 14, SOFA score: 3, temperature: 37.6°C, RDW: 14%, serum sodium: 140 mmol/L; Serum potassium: 3.8 mmol/L; oxygen saturation: 96%. The predicted probability of occurrence in this patient was 9.181%, which indicates that the patient had a low risk of in-hospital mortality (Figure 6B) .

**Figure 6.**
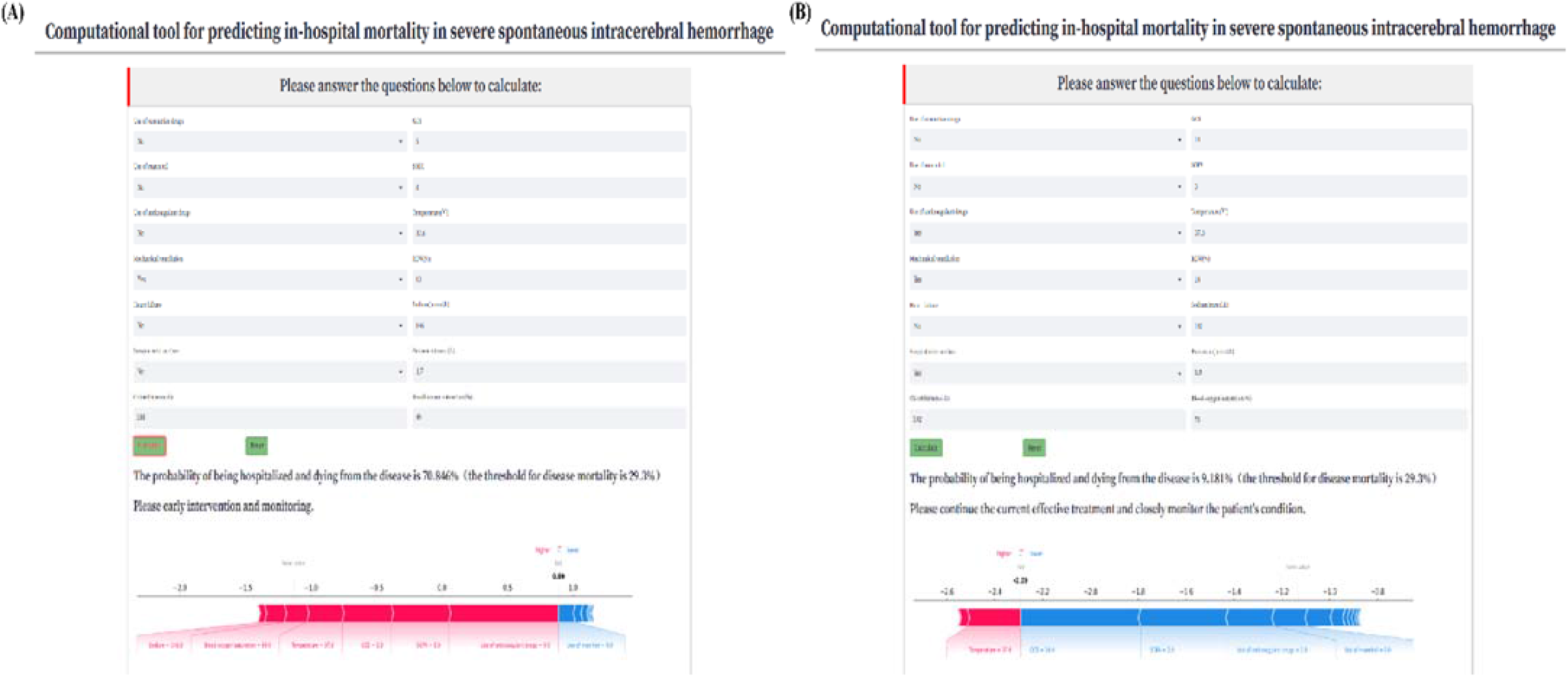
Cases of website usage. Enter input values to determine the prognosis for sICH, and show the contribution of the variable shap value to the prediction. (A) Case 1 has a high probability of in-hospital death; (B) Case 2 has a high probability of a better prognosis.

## Discussion

In this retrospective study, we developed and validated a clinical feature-based machine learning algorithm for predicting in-hospital mortality in critically ill patients with sICH based on the public database MIMIC-IV and the Zhejiang Hospital database. Among the tested models, the XGBOOST model demonstrated the highest prediction performance. Through advanced machine learning techniques, we successfully identified several key clinical features strongly associated with in-hospital mortality, including GCS score, SOFA score, use of mannitol medication, use of anticoagulant medication, vital signs, serum electrolytes, RDW, among others. These findings are significant and warrant further investigation. Additionally, we have created an easy-to-use web-based calculator to assist clinicians in making informed decisions regarding further treatment.

Among various types of strokes, cerebral hemorrhage is characterized by a relatively high in-hospital mortality rate, especially in patients admitted to ICU. The in-hospital mortality rate of patients varies based on both the location and volume of the hematoma. Previous studies have reported an early mortality rate of 40% and a long-term mortality rate as high as 60% for sICH patients ^13–15^.Marika Fallenius et al. analyzed patients admitted to the ICU with severe cerebral hemorrhage and found a mortality rate of 42% for supratentorial sICH patients and 49% for infratentorial sICH patients ^16^. Additionally, researchers investigated a 30-day mortality rate of up to 54% for patients with severe sICH in the southern region of Spain, and this rate increased to 60% for patients with hematoma volumes exceeding 30 ml ^17^. The mortality rate observed in our study was lower compared to the case-fatality rates reported in previous studies. This difference may be attributed to our exclusion of patients admitted for less than one day or those automatically discharged, as well as differences in medical conditions. Nevertheless, identifying patients at high risk of in-hospital mortality in the ICU remains challenging for clinicians. Thus, there is an urgent need for the development and widespread dissemination of reliable predictive models. Such models would play a crucial role in identifying high-risk patients and enabling timely and effective interventions to enhance their prognosis.

Currently, research in the field of predictive modeling of diseases is rapidly advancing, thanks to the increasing applicability and effectiveness of supervised machine learning algorithms ^18,19^. Several well-known supervised learning classifiers, such as KNN, support vector machines, Random Forests, convolutional neural networks, XGBoost, Light gradient boosting, and AdaBoost, have found progressive application in clinical practice ^20–22^. Through the application of ML classification, research has demonstrated that ML-assisted decision support models offer advantages over traditional linear regression models. In this study, we employed five distinct ML methods to develop predictive models. The performance evaluation of these algorithms was based on six common metrics (AUC, F1 score, accuracy, sensitivity, specificity and Youden Index). Notably, the results unequivocally indicate that the XGBoost model exhibits the most superior performance and predictive stability, which contrasts with previous findings favoring the Random Forest model ^23^. XGBoost is an efficient, flexible, and scalable ML algorithm, renowned for its classification capabilities. To mitigate overfitting and optimize its performance, XGBoost employs techniques such as improved subsampling rates, learning rates, and maximum tree depth control ^24^. Zhu et al. evaluated data from ICU patients who were intubated due to respiratory failure and received mechanical ventilation. They utilized seven learning algorithms to predict in-hospital mortality, with XGBoost demonstrating the best overall performance^25^. Similarly, Hu et al. incorporated data from 8817 sepsis patients into seven models to predict in-hospital mortality, and they also found that the XGBoost model exhibited the most effective predictive ability^26^.

Despite the success of algorithms in this field, one of the current challenges lies in the need to interpret the “black box” of ML. Thus, we utilized the visualization function in SHAP to identify the impact of specific variable values on the model output. SHAP is a game-theoretic technique developed by Lundberg and Lee, effectively addressing the black-box nature of ML models by providing consistent interpretability. By employing an advanced ML approach, we identified the top 10 predictors in the rankings of feature importance, which included GCS, SOFA, anticoagulant medication use, mannitol medication use, oxygen saturation, temperature, sodium, potassium, RDW, and mechanical ventilation The GCS is a widely used scale for assessing the level of consciousness, with scores ranging from 3 to 15. Previous studies have consistently demonstrated the importance of the GCS score in evaluating the severity of neurological disorders ^23,27,28^. The SOFA score serves as a valuable tool for quantifying the extent of organ dysfunction or failure at the point of ICU admission and has found widespread application in predicting in-hospital mortality in this setting ^29–31^. It has been observed that the SOFA score exhibits superior predictive performance compared to other scoring systems when it comes to infection-related in-hospital mortality in ICU patients ^32^. The use of anticoagulants in patients with cerebral hemorrhage and the timing of anticoagulant use remain controversial, and some studies have suggested that anticoagulants have a positive effect on patient prognosis. This might be because the use of anticoagulants in critically ill patients reduces complications such as thromboembolism and does not significantly increase bleeding complications ^33^.

Currently, the primary non-surgical treatment for cerebral hemorrhage involves the use of drugs like mannitol to reduce intracranial pressure. However, our study revealed that the use of mannitol may lead to a poor prognosis for patients with this disease. According to current guidelines, hypertonic saline demonstrates superior efficacy in managing cerebral edema associated with cerebral hemorrhage compared to mannitol ^34^. Moreover, prior research has established that electrolyte disturbances represent an independent risk factor for an unfavorable prognosis in stroke patients^35–37^. Lastly, to our surprise, the study indicates that in critically ill patients, surgical treatment may not hold significant importance. It is plausible that conservative pharmacological treatment has a more positive effect on the prognosis in these patients compared to secondary trauma.

This study holds significant clinical and methodological implications. Firstly, we implemented an external validation set to mitigate the risk of model overfitting. Secondly, the model was developed using readily available data collected within 24 hours of patient admission, enabling early and accurate mortality prediction. This provides clinicians with more time to adjust treatment strategies accordingly. Thirdly, the study sheds light on previously overlooked factors, such as anticoagulant use and RDW, which are now identifiable. Integrating these factors with machine learning methods enhances the predictive performance significantly. Lastly, to facilitate bedside use by clinicians, a user-friendly calculator based on the model was developed.

However, our study has several limitations that need to be acknowledged. Firstly, it was a retrospective and observational study, which may introduce certain research biases. Secondly, as our study was focused on patients with sICH, we did not include information on radiologic variables, such as hematoma volume or location, which could potentially provide additional insights into the disease.

## Conclusion

The XGBoost model demonstrated superior performance compared to the logistic regression (LR), K-nearest neighbors (KNN), Random Forest (RF), and adaptive boosting (AdaBoost) models in predicting short-term mortality among sICH patients. Our findings indicate that factors such as GCS, SOFA score, mannitol use, anticoagulant use, oxygen saturation, time of ICU admission, temperature, serum sodium, mechanical ventilation, and serum potassium are strongly associated with in-hospital mortality in sICH patients. This newly developed risk model is expected to serve as a convenient tool for risk stratification.

## Data Availability

The datasets generated during and/or analyzed during the current study are available from the corresponding author on reasonable request.

## Acknowledgments

We would like to thank all the participants and investigators that took part in this study.

## Sources of Funding

This work was supported by the grants from Special Support Program for High Level Talents of Zhejiang Province(2022R52038), Key Research and Development Project of Zhejiang Provincial Department of Science and Technology(2021C03105), Natural Science Foundation of Zhejiang Province (LY21H090008) and Medical Health Science and Technology Project of Zhejiang Provincial Health Commission (WKJ-ZJ-2340).

## Disclosures

All authors declare no conflicts of interest.

## Notes

### Competing Interest Statement

The authors have declared no competing interest.

### Author Declarations

The study protocol was approved by the Ethics Committee of Zhejiang Hospital (No. 2023 Pro-examination (58K)).

## Reference

1. Qureshi AI, Tuhrim S, Broderick JP, Batjer HH, Hondo H, Hanley DF. Spontaneous intracerebral hemorrhage. N Engl J Med. 2001;344:1450–1460.

2. Aronowski J, Zhao X. Molecular Pathophysiology of Cerebral Hemorrhage: Secondary Brain Injury. Stroke. 2011;42:1781–1786.

3. Wang J. Preclinical and clinical research on inflammation after intracerebral hemorrhage. Prog Neurobiol. 2010;92:463–477.

4. Esteva A, Kuprel B, Novoa RA, Ko J, Swetter SM, Blau HM, Thrun S. Dermatologist-level classification of skin cancer with deep neural networks. Nature. 2017;542:115–118.

5. Ambale-Venkatesh B, Yang X, Wu CO, Liu K, Hundley WG, McClelland R, Gomes AS, Folsom AR, Shea S, Guallar E, Bluemke DA, Lima JAC. Cardiovascular Event Prediction by Machine Learning: The Multi-Ethnic Study of Atherosclerosis. Circ Res. 2017;121:1092–1101.

6. Nomura A, Noguchi M, Kometani M, Furukawa K, Yoneda T. Artificial Intelligence in Current Diabetes Management and Prediction. Curr Diab Rep. 2021;21:61.

7. Huang B, Liang D, Zou R, Yu X, Dan G, Huang H, Liu H, Liu Y. Mortality prediction for patients with acute respiratory distress syndrome based on machine learning: a population-based study. Ann Transl Med. 2021;9:794.

8. Huang T, Le D, Yuan L, Xu S, Peng X. Machine learning for prediction of in-hospital mortality in lung cancer patients admitted to intensive care unit. PloS One. 2023;18:e0280606.

9. Raita Y, Goto T, Faridi MK, Brown DFM, Camargo CA, Hasegawa K. Emergency department triage prediction of clinical outcomes using machine learning models. Crit Care Lond Engl. 2019;23:64.

10. Manz CR, Chen J, Liu M, Chivers C, Regli SH, Braun J, Draugelis M, Hanson CW, Shulman LN, Schuchter LM, O’Connor N, Bekelman JE, Patel MS, Parikh RB. Validation of a Machine Learning Algorithm to Predict 180-Day Mortality for Outpatients With Cancer. JAMA Oncol. 2020;6:1723–1730.

11. Bunney G, Murphy J, Colton K, Wang H, Shin HJ, Faigle R, Naidech AM. Predicting Early Seizures After Intracerebral Hemorrhage with Machine Learning. Neurocrit Care. 2022;37:322–327.

12. Rusche T, Wasserthal J, Breit H-C, Fischer U, Guzman R, Fiehler J, Psychogios M-N, Sporns PB. Machine Learning for Onset Prediction of Patients with Intracerebral Hemorrhage. J Clin Med. 2023;12:2631.

13. Van Asch CJ, Luitse MJ, Rinkel GJ, Van Der Tweel I, Algra A, Klijn CJ. Incidence, case fatality, and functional outcome of intracerebral haemorrhage over time, according to age, sex, and ethnic origin: a systematic review and meta-analysis. Lancet Neurol. 2010;9:167–176.

14. Sacco S, Marini C, Toni D, Olivieri L, Carolei A. Incidence and 10-Year Survival of Intracerebral Hemorrhage in a Population-Based Registry. Stroke. 2009;40:394–399.

15. Poon MTC, Fonville AF, Al-Shahi Salman R. Long-term prognosis after intracerebral haemorrhage: systematic review and meta-analysis. J Neurol Neurosurg Psychiatry. 2014;85:660–667.

16. Fallenius M, Skrifvars MB, Reinikainen M, Bendel S, Curtze S, Sibolt G, Martinez-Majander N, Raj R. Spontaneous Intracerebral Hemorrhage. Stroke. 2019;50:2336–2343.

17. Rodríguez-Fernández S, Castillo-Lorente E, Guerrero-Lopez F, Rodríguez-Rubio D, Aguilar-Alonso E, Lafuente-Baraza J, Gómez-Jiménez FJ, Mora-Ordóñez J, Rivera-López R, Arias-Verdú MD, Quesada-García G, Arráez-Sánchez MÁ, Rivera-Fernández R. Validation of the ICH score in patients with spontaneous intracerebral haemorrhage admitted to the intensive care unit in Southern Spain. BMJ Open. 2018;8:e021719.

18. Sidey-Gibbons JAM, Sidey-Gibbons CJ. Machine learning in medicine: a practical introduction. BMC Med Res Methodol. 2019;19:64.

19. Hueman M, Wang H, Liu Z, Henson D, Nguyen C, Park D, Sheng L, Chen D. Expanding TNM for lung cancer through machine learning. Thorac Cancer. 2021;12:1423–1430.

20. Uddin S, Khan A, Hossain ME, Moni MA. Comparing different supervised machine learning algorithms for disease prediction. BMC Med Inform Decis Mak. 2019;19:281.

21. Dinh A, Miertschin S, Young A, Mohanty SD. A data-driven approach to predicting diabetes and cardiovascular disease with machine learning. BMC Med Inform Decis Mak. 2019;19:211.

22. Osman MH, Mohamed RH, Sarhan HM, Park EJ, Baik SH, Lee KY, Kang J. Machine Learning Model for Predicting Postoperative Survival of Patients with Colorectal Cancer. Cancer Res Treat. 2022;54:517–524.

23. Nie X, Cai Y, Liu J, Liu X, Zhao J, Yang Z, Wen M, Liu L. Mortality Prediction in Cerebral Hemorrhage Patients Using Machine Learning Algorithms in Intensive Care Units. Front Neurol. 2020;11:610531.

24. Xu Y, Han D, Huang T, Zhang X, Lu H, Shen S, Lyu J, Wang H. Predicting ICU Mortality in Rheumatic Heart Disease: Comparison of XGBoost and Logistic Regression. Front Cardiovasc Med. 2022;9:847206.

25. Zhu Y, Zhang J, Wang G, Yao R, Ren C, Chen G, Jin X, Guo J, Liu S, Zheng H, Chen Y, Guo Q, Li L, Du B, Xi X, Li W, Huang H, Li Y, Yu Q. Machine Learning Prediction Models for Mechanically Ventilated Patients: Analyses of the MIMIC-III Database. Front Med. 2021;8:662340.

26. Hu C, Li L, Huang W, Wu T, Xu Q, Liu J, Hu B. Interpretable Machine Learning for Early Prediction of Prognosis in Sepsis: A Discovery and Validation Study. Infect Dis Ther. 2022;11:1117–1132.

27. Teasdale G, Jennett B. Assessment of coma and impaired consciousness. A practical scale. Lancet Lond Engl. 1974;2:81–84.

28. Agrawal N, Iyer SS, Patil V, Kulkarni S, Shah JN, Jedge P. Comparison of admission GCS score to admission GCS-P and FOUR scores for prediction of outcomes among patients with traumatic brain injury in the intensive care unit in India. Acute Crit Care. 2023;38:226–233.

29. Ferreira FL. Serial Evaluation of the SOFA Score to Predict Outcome in Critically Ill Patients. JAMA. 2001;286:1754.

30. Vincent J-L, Moreno R, Takala J, Willatts S, De Mendonça A, Bruining H, Reinhart CK, Suter PM, Thijs LG. The SOFA (Sepsis-related Organ Failure Assessment) score to describe organ dysfunction/failure: On behalf of the Working Group on Sepsis-Related Problems of the European Society of Intensive Care Medicine (see contributors to the project in the appendix). Intensive Care Med. 1996;22:707–710.

31. Cárdenas-Turanzas M, Ensor J, Wakefield C, Zhang K, Wallace SK, Price KJ, Nates JL. Cross-validation of a Sequential Organ Failure Assessment score–based model to predict mortality in patients with cancer admitted to the intensive care unit. J Crit Care. 2012;27:673–680.

32. Raith EP, Udy AA, Bailey M, McGloughlin S, MacIsaac C, Bellomo R, Pilcher DV, for the Australian and New Zealand Intensive Care Society (ANZICS) Centre for Outcomes and Resource Evaluation (CORE). Prognostic Accuracy of the SOFA Score, SIRS Criteria, and qSOFA Score for In-Hospital Mortality Among Adults With Suspected Infection Admitted to the Intensive Care Unit. JAMA. 2017;317:290.

33. Kuramatsu JB, Huttner HB. Management of oral anticoagulation after intracerebral hemorrhage. Int J Stroke Off J Int Stroke Soc. 2019;14:238–246.

34. Cook AM, Morgan Jones G, Hawryluk GWJ, Mailloux P, McLaughlin D, Papangelou A, Samuel S, Tokumaru S, Venkatasubramanian C, Zacko C, Zimmermann LL, Hirsch K, Shutter L. Guidelines for the Acute Treatment of Cerebral Edema in Neurocritical Care Patients. Neurocrit Care. 2020;32:647–666.

35. Bales J, Cho S, Tran TK, Korab GA, Khandelwal N, Spiekerman CF, Joffe AM. The Effect of Hyponatremia and Sodium Variability on Outcomes in Adults with Aneurysmal Subarachnoid Hemorrhage. World Neurosurg. 2016;96:340–349.

36. Matano F, Fujiki Y, Mizunari T, Koketsu K, Tamaki T, Murai Y, Yokota H, Morita A. Serum Glucose and Potassium Ratio as Risk Factors for Cerebral Vasospasm after Aneurysmal Subarachnoid Hemorrhage. J Stroke Cerebrovasc Dis Off J Natl Stroke Assoc. 2019;28:1951–1957.

37. Sadan O, Samuels O, Asbury WH, Hanfelt JJ, Singbartl K. Low-chloride versus high-chloride hypertonic solution for the treatment of subarachnoid hemorrhage-related complications (The ACETatE trial): study protocol for a pilot randomized controlled trial. Trials. 2018;19:628.

